# Targeted saliva multi-omics is a reliable, non-invasive method to capture physiological stress and recovery

**DOI:** 10.64898/2026.01.28.26345066

**Authors:** Charlotte Wenzel, Bürkan Kalaycik, Annika Billig, Sina Trebing, Niklas Joisten, Mathias Kolodziej, Markus Braun, Lars Lippelt, Alexander Gerharz, Marlon Millard, Oliver Wieder, Karin Kipper, Adam Iebed, Andreas Groll, David Walzik, Philipp Zimmer

## Abstract

Determining physiological stress at high resolution is crucial across diverse settings to enable informed decision-making in the context of health and disease. Saliva-based targeted multi-omics testing provides a powerful, non-invasive method to quantify physiological stress and circadian dynamics at high-frequency. In a laboratory crossover trial with 24-hour sampling comprising 413 saliva samples, we demonstrate high analytical reliability, distinct molecular individuality, and robust acute and delayed responses to physical exercise across proteins, metabolites, and lipids. Moreover, we present the most comprehensive existing dataset describing 24-hour molecular kinetics across these three omics layers. Leveraging this controlled setting, we applied machine learning to single-timepoint saliva samples to accurately predict recent physical exercise both immediately after and 24 hours later. Next, we translated this analytical framework to a real-world longitudinal setting of elite football players monitored over 16 months, comprising over 12,000 saliva samples. Despite increased biological and contextual variability, the model retained robust discrimination between exercise and rest on the following day. Based on prediction probabilities, we introduce a saliva-based internal strain metric, that captures internal load and can be harnessed to monitor physical exercise and recovery. Model robustness was further supported through out-of-sample validation using previously unseen observations. Our findings demonstrate that saliva-based targeted multi-omics reliably captures physical exercise and recovery states in both laboratory and real-world environments, providing a scalable framework for monitoring physical performance. This non-invasive approach holds broad potential for physiological monitoring and can serve as a blueprint for health- and disease-related contexts.

## INTRODUCTION

Physiological stress as provoked by external (e.g. physical exercise, drug intake) or internal stimuli (e.g. pathological conditions) is a dynamic and complex state that involves the integrated response of multiple biological systems^1,2^. Accurate measurement of these dynamics with high temporal resolution is crucial in various fields, including personalised health monitoring, disease prevention, and performance optimisation, as it enables informed, context-specific decision-making. However, individualised, time-resolved tools for quantifying physiological stress and recovery in real-world settings remain elusive to date.

High-frequency non-invasive biomarker quantification has therefore gained increasing attention in settings ranging from elite sports^3^ to healthcare and routine clinical diagnostics^4^. Despite the broad impact of digital biomarkers such as step counts and GPS-data^3^, few non-invasive measures display an individual’s physiological or health state. Established examples include heart rate, blood pressure, and measures of body composition^5,6^. However, more detailed biological outcomes are almost exclusively derived from venous or capillary blood, limiting their applicability for day-to-day use and high-frequency monitoring. Venous blood collection requires trained staff, and capillary point-of-care analyses are usually limited to single outcomes such as glucose or lactate^7^. Most established biomarkers also reflect isolated biological processes rather than an individual’s integrated physiological state, constraining their ability to capture dynamic processes such as physiological stress, recovery, or circadian modulation.

Together, these limitations highlight the need for non-invasive biomarkers that capture integrated physiological states with high temporal resolution. In this context, saliva depicts a valuable biofluid since it contains thousands of proteins, metabolites, and lipids, which reflect an individual’s physiology^8,9^. Initial evidence suggests that saliva testing may serve as a powerful non-invasive alternative to blood-based analyses for assessing acute responses to physical exercise^8^. However, to date, comprehensive, high-frequency quantification of saliva-based analytes is lacking and restricted to either small numbers of analytes (e.g., cortisol)^10,11^, few measurement time points^12^, or, in the case of comprehensive analyses, to case reports^13^.

To address these limitations, we conducted high-frequency targeted multi-omics analyses (i.e., proteomics, metabolomics, and lipidomics) of saliva samples. Our analyte panel was designed based on the exercise responsiveness and quantification reliability of saliva analytes. In a randomised controlled crossover trial, we collected saliva samples over a 24-hour period following an acute exercise bout. This dense sampling enabled us to assess test-retest and inter-day reliability, as well as inter-individual variability. We further provide a comprehensive characterisation of 24-hour circadian dynamics across three omics layers. The obtained data also allowed evaluation of acute and delayed molecular responses to exercise. Building upon these findings, we applied different machine learning approaches to single-timepoint saliva samples, demonstrating that targeted multi-omics profiles can reliably predict recent physical exercise. This modelling framework was then transferred to a real-world longitudinal cohort of elite football players. From this real-world setting, we derive and validate a new saliva-based metric, which captures internal load following recent physical exercise and can be harnessed for recovery monitoring.

## RESULTS

### Targeted saliva multi-omics demonstrates high reliability and molecular individuality in a standardised laboratory setting

We investigated a controlled laboratory cohort and a longitudinal real-world cohort (Fig. 1a). Twelve healthy, overnight-fasted females (age [years]: 15.17 ± 1.03, BMI [kg/m^2^]: 20.2 ± 2.1; mean ± standard deviation; supplementary Table 1) were included in the final analysis of a randomised crossover laboratory study with two testing days. On one day, participants underwent maximal cardiopulmonary exercise testing (CPET) on a cycle ergometer (V O_2_peak [ml·kg^−1^·min^−1^]: 44.17 ± 5.39) between 7:00 and 11:00 a.m., followed by a verification phase (VP), while the other day served as a passive control. Saliva samples were collected at identical time points on both days: twice at baseline, immediately after CPET, after the VP, and 15, 30, and 60 minutes after the VP. Following the last supervised sample, participants consumed a standardised meal and self-collected saliva samples every two hours for 24 hours, resulting in a total of 413 collected samples.

**Fig. 1.**
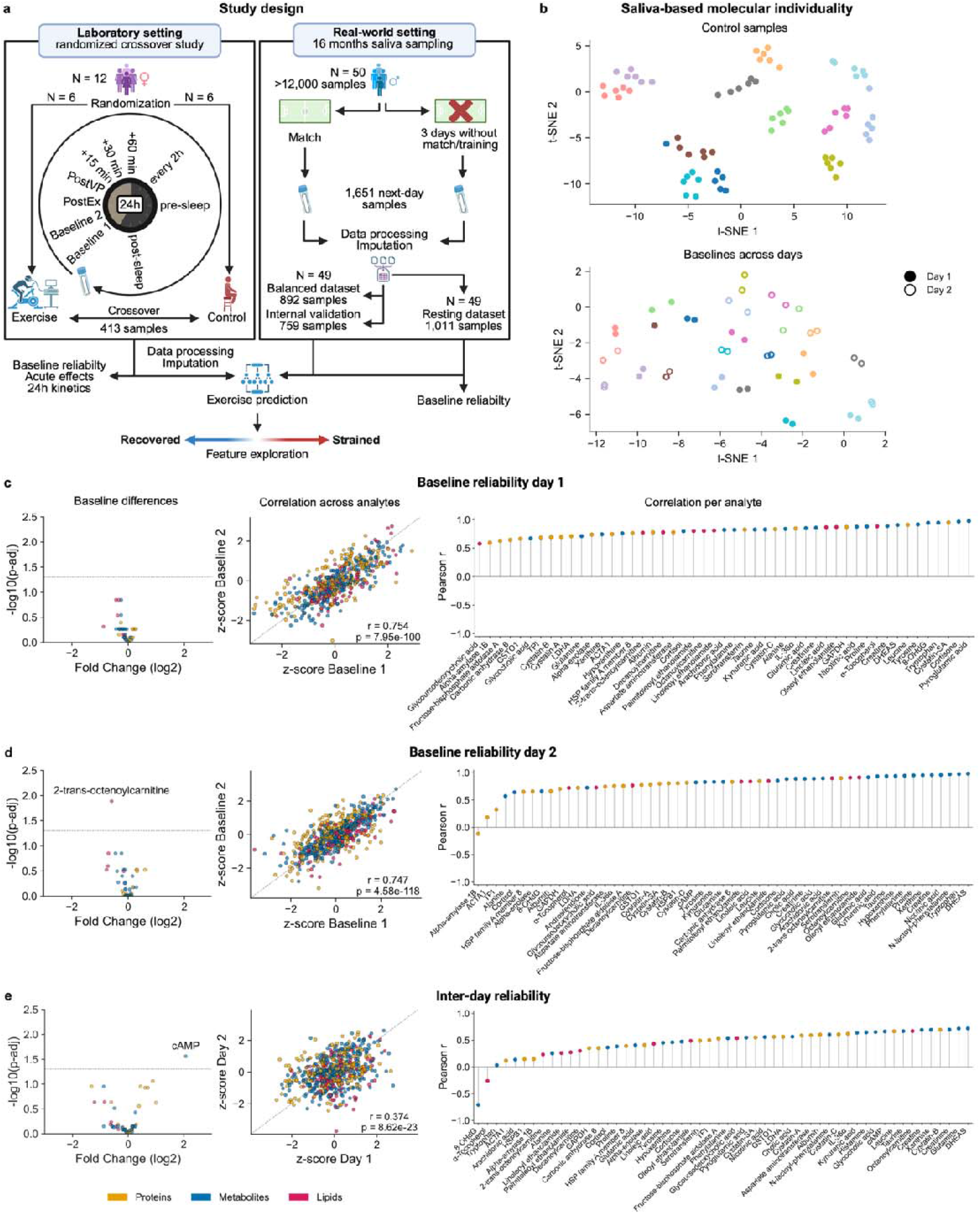
Study design, molecular individuality, and baseline reliability of targeted saliva multi-omics. **a**, Study design and analysis workflow. **b**, t-distributed stochastic neighbour embeddings (t-SNE) indicate high molecular individuality, consistent with saliva-based fingerprints. Samples are coloured by participant ID. **c-e**, Baseline reliability of saliva analytes on day 1 (**c**), day 2 (**d**), and across days (**e**). Reliability was assessed via two-sided, Benjamini-Hochberg-corrected, paired Student’s t-tests (left), overall correlation analyses (middle), and per-analyte correlations (right), respectively. N = 12 with 49 - 55 analytes per sample for all analyses.

By conducting dimensionality reduction analyses with passive control samples we observed high molecular individuality, indicative of saliva-based “fingerprints”, thus confirming strong inter-individual variation in analyte patterns^6,11^. Interestingly, these fingerprints were retained even when baseline samples from both intervention days were included in the analysis, suggesting temporal stability of saliva profiles and the possibility of long-term saliva-based molecular phenotyping (Fig. 1b). To assess the reliability of our targeted multi-omics approach, we compared baseline measurements within and between intervention days by testing analyte-wise concentration differences, complemented by global correlation analyses across all analytes and per-analyte correlations (Fig. 1c-e). Only one lipid (2-trans-octenoylcarnitine) differed between consecutive baseline measurements. Overall correlation between successive baseline samples showed high intra-day reliability for day one (r = 0.75; p = 7.95 × 10^-100^) and for day two (r = 0.75; p = 4.58 × 10^-118^). Similarly, per-analyte correlations of consecutive baseline samples were moderate to high for most analytes on both days (Fig. 1c,d). Inter-day comparisons of baseline samples also demonstrated good reproducibility with only one metabolite (Cyclic adenosine monophosphate) differing significantly between days. Global correlation across all analytes was slightly lower (r = 0.37; p = 8.62 × 10^-23^), likely reflecting increased biological variability between days rather than analytical noise. Of all quantified analytes, most retained moderate to strong positive correlations across days (Fig. 1e). Overall, these results confirm the reliability of our method and suggest high molecular individuality in saliva profiles, which served as crucial prerequisite and first insight into our new approach.

### High-frequency saliva sampling depicts circadian rhythms across different omics layers

Next, we examined 24-hour kinetics of all analytes quantified in control samples to assess saliva-based circadian fluctuations (Fig. 2a-c, supplementary Table 2). Well-established analytes such as Cortisol and Melatonin showed expected rhythms^14–16^, however, we were also able to replicate circadian profiles for analytes like Hypoxanthine and the bile acid-conjugate Glycoursodeoxycholic acid, which were previously reported only in blood^17,18^. Interestingly, we also observed circadian fluctuations for Taurine, which itself was shown to regulate circadian rhythms^19^, and for N-lactoyl-phenylalanine, which is highly exercise-responsive^20^. Kynurenic acid, a metabolite with immuno- and neuromodulatory properties^21^, also showed slight circadian variation, which is a novel observation for saliva. Among salivary proteins, we observed contrasting temporal patterns. Notably, Alpha-amylase 1B exhibited a 24-hour pattern that was opposite to most other proteins, including Alpha-enolase (Fig. 2a). This deviation might reflect different regulatory mechanisms, whereby certain proteins are predominantly controlled by the activity of the sympathetic nervous system, while others are rather affected by local metabolic processes^22^. In contrast, a subset of analytes, such as Heat shock protein β-1 (HSPB1), Glutamic acid or Decanoylcarnitine, showed no 24-hour rhythms, which might be explained through unresponsiveness of these analytes to endogenous timing mechanisms. Of note, one strength of the targeted approach applied here is the determination of absolute concentrations in saliva-based analytes over a 24-hour cycle (supplementary Table 2), which provides a solid basis for establishing individual reference ranges^23^. Importantly, several of these circadian patterns have been described primarily in blood and, when reported in saliva, have not previously been captured at such high temporal resolution^24^. Together, this foundation enables the detection of (patho)physiological disruptions to homeostasis, for example in the context of exercise or disease.

**Fig. 2.**
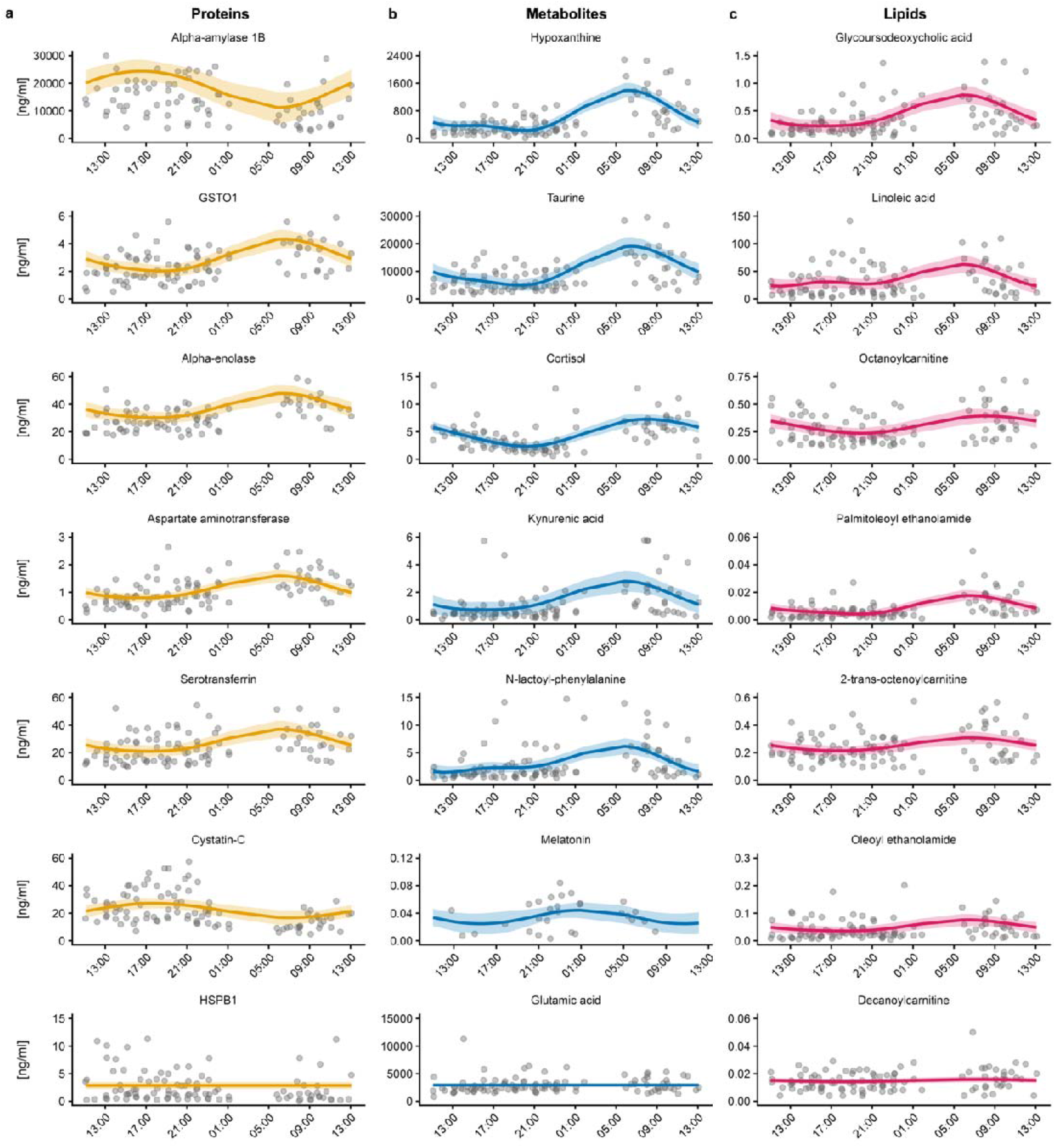
24-hour kinetics of selected saliva analytes. **a-c**, Kinetics of proteins (**a**), metabolites (**b**), and lipids (**c**) over a 24-hour cycle. Temporal trends were modelled using Bayesian additive models. Mean model prediction and 95% confidence intervals are displayed alongside raw data points. N = 12 with 53 analytes per sample for all analyses. See also supplementary Table 2.

### Identification of immediate and delayed exercise responses of saliva-based analytes

Beyond characterising the 24-hour resting profiles, we examined the acute response of saliva-based analytes to a single exercise bout. Lipids exhibited the greatest variability across timepoints and conditions (83.61%; average coefficient of variation over time points), followed by metabolites (70.25%) and proteins (53.38%) (Fig. 3a). Linear mixed models (LMM) identified 13 analytes with significant interaction effects between exercise and control, thus suggesting a novel application of these analytes as saliva-based exercise biomarkers (Fig. 3b). While many metabolites decreased in the morning hours of the control day - a pattern that is consistent with circadian regulation - most of our newly identified exercise biomarkers increased immediately or shortly after exercise (Fig. 3c). Hierarchical clustering of exercise responses revealed coherent clusters of molecular analytes that exhibited similar time-course response patterns under exercise and control conditions (Fig. 3d). These clusters highlight groups of analytes that respond in a coordinated manner to physical exercise and recovery, suggesting common regulation or involvement in related biological processes. In line with previous detailed characterisations of the molecular response to exercise^25^, this highlights similar patterns in saliva.

**Fig. 3.**
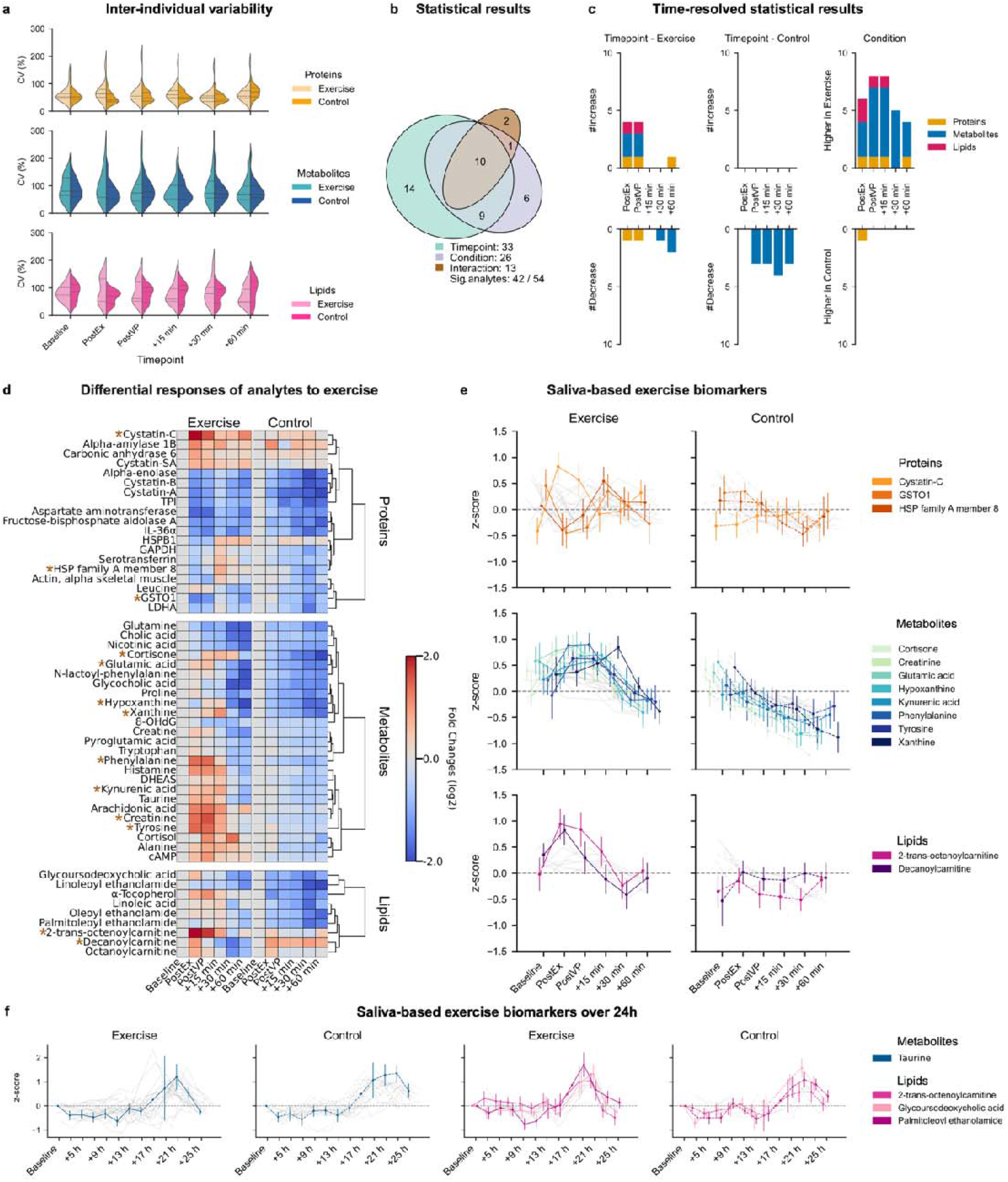
Targeted saliva multi-omics identifies exercise biomarkers. **a**, Inter-individual variability expressed as coefficient of variation across conditions and timepoints separated by omics layer. The width of each violin plot indicates the distribution density, while the inner lines mark the 25^th^, 50^th^ (median) and 75^th^ percentiles. **b**, Overview of statistical results derived from linear mixed models (LMM) including fixed effects for timepoint, condition, and the interaction between both, and a random intercept for participant ID. **c**, Post-hoc analyses of analytes showing significant interaction effects (= exercise biomarkers). Two-sided, Bonferroni-corrected, paired Student’s t-tests were performed to evaluate differences over time in each condition as well as differences between the two conditions at each timepoint. **d**, Temporal regulation of saliva analytes in response to exercise vs. control. Hierarchical clustering was applied to identify analytes displaying similar kinetics. Exercise biomarkers are marked with an asterisk. **e**, Exercise kinetics of analytes demonstrating interaction effects compared to control (mean ± SEM). Grey lines indicate analytes that did not demonstrate interaction effects. N = 12 with 54 analytes. f, Exercise kinetics of analytes demonstrating interaction effects over a 24-hour cycle (= 24-hour exercise biomarkers). Grey lines indicate analytes that did not demonstrate interaction effects. N = 12 with 52 analytes.

In detail, we observed distinct temporal dynamics of the 13 exercise biomarkers in response to exercise versus control (Fig. 3e). On the exercise day, most molecules showed an initial increase immediately after physical exercise, followed by a rapid decrease to or below baseline levels within 30 minutes after. Lipids subsequently rose again and returned to baseline by 60 minutes, whereas metabolites generally continued to decrease. Among proteins, Cystatin-C remained elevated above baseline 60 minutes after exercise, while heat shock protein (HSP) family A, member 8 and Glutathione S-transferase omega 1 (GSTO1) generally exhibited opposite time-course dynamics. The temporal trends on the control day were comparable within each omics layer.

Over the 24-hour period, LMM identified four analytes (Taurine, 2-trans-octenoylcarnitine, Glycoursodeoxycholic acid, Palmitoleoyl ethanolamide) that demonstrated differential regulation between the exercise and control condition (Fig. 3f). Notably, these analytes showed a distinct peak between 17 and 25 hours after exercise and control, suggesting a potential role as exercise biomarkers beyond the acute recovery window.

Overall, our identification of exercise-responsive biomarkers depicts a crucial step forward for biomarker testing in applied exercise settings, where practical usability has been limited to analytes like creatine kinase (CK)^26^or C-reactive protein (CRP)^27^ or a long time.

### Single timepoint saliva samples capture acute and delayed responses to physical exercise

We next used supervised prediction analyses to evaluate whether a single-timepoint saliva sample can be used in a binary classification task to reliably distinguish between exercise and control immediately after exercise and then assessed whether saliva samples still contained similar biological signals 24 hours later. Using a cross-validated model selection framework, the best-performing model for samples collected immediately after exercise (random forest-based feature pre-selection followed by ridge-penalised logistic regression) achieved a predictive receiver operating characteristic area under the curve (ROC-AUC) value of 0.95. The Bernoulli likelihood was 0.68 (Fig. 4a), reflecting that the model assigned high probabilities to the correct class labels on average. Twenty features were retained in the final model. Notably, nine of the previously identified exercise biomarkers (Fig. 3e) were also among the most influential features in the ridge-based classification framework (Fig. 4b). Cystatin-C showed the largest standardized regression coefficient, followed by 2-trans-octenoylcarnitine, consistent with their pronounced early concentration changes immediately after exercise (Fig. 3e).

**Fig. 4.**
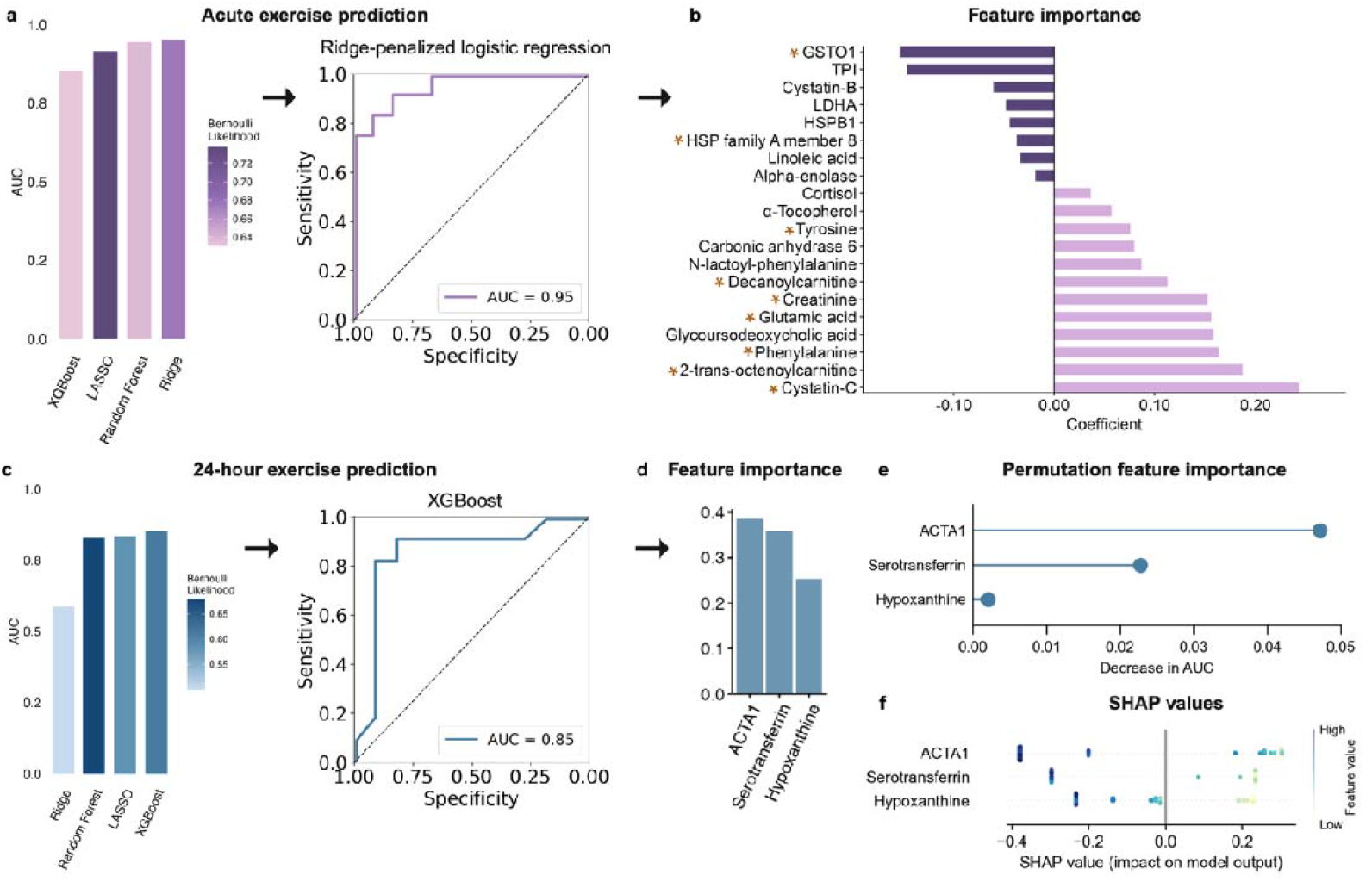
Targeted saliva multi-omics captures physical exercise acutely and after 24 hours. **a**, Comparison of the predictive receiver operating characteristic area under the curve (ROC-AUC) values across four models for classification of exercise vs. control immediately after exercise, and ROC curve of the best-performing classifier (ridge-penalised logistic regression). Bernoulli likelihood is color-coded. **b**, Feature importance shown as coefficient estimates from ridge-penalised logistic regression (standardized features). Exercise biomarkers are marked with an asterisk. **c**, ROC-AUC values across four models for classification of exercise vs. control 24 hours after exercise, and ROC curve of the best-performing classifier (XGBoost). Bernoulli likelihood is color-coded. **d**, Gain-based feature importance of XGBoost **e**, permutation feature importance of XGBoost **f**, and SHapley Additive exPlanations (SHAP) values of XGBoost.

As expected, predictive performance decreased 24 hours after exercise due to increased interindividual variability. Nevertheless, the combination of LASSO-based feature pre-selection and extreme gradient-boosted decision trees (XGBoost) classifier achieved the best performance among all evaluated models, with predictive ROC-AUC and Bernoulli likelihood values of 0.85 and 0.61, respectively (Fig. 4c). Interestingly, this final model was based exclusively on three features and still retained considerable discriminatory power, suggesting that large amounts of biological information regarding exercise exposure the day prior are contained in these three saliva-based analytes. Gain-based feature importance analyses identified Actin, alpha-skeletal muscle (ACTA1) as the dominant feature, followed by Serotransferrin and Hypoxanthine (Fig. 4d), which is consistent with their biological functions. ACTA1, a structural muscle protein, can be released from skeletal muscle in response to exercise-induced muscle damage following mechanical stress^18,19^, and Serotransferrin, a key iron transport protein, may reflect exercise-induced changes in iron handling, oxygen transport, and inflammatory signalling^20^, thus also linking to exercise. Hypoxanthine, a purine degradation product, reflects increased ATP turnover and metabolic stress^21^. Permutation analyses revealed that randomly permuting ACTA1 reduced the ROC-AUC value by approximately 0.05 (Fig. 4e), underscoring its crucial contribution to model performance. Interpretation of Shapley’s Additive Explanations (SHAP) yielded a consistent direction for all three features (Fig. 4f) with lower feature values shifting the predictions towards the exercise condition, which is in line with the observed 24-hour kinetics.

In summary, the laboratory results demonstrate that, under controlled conditions, a single-timepoint saliva sample provides robust discrimination between exercise and control both immediately and 24 hours after physical exercise. This creates a malleable foundation for the introduction of saliva-based targeted multi-omics tools to monitor exercise strain and recovery in real-world settings.

### Real-world sampling confirms molecular individuality and high reliability of targeted saliva multi-omics

To assess whether the molecular individuality and reliability identified under controlled laboratory conditions can be transferred to a real-world setting, we analysed longitudinal saliva samples collected from elite football players throughout 16 months (Fig. 1a). From a total of over 12,000 samples, our final analysis cohort comprised 49 male elite athletes, from whom a total of 1,651 saliva samples were selected based on predefined exercise and rest criteria: one day after a match (exercise condition) or after at least three consecutive days without training/match (control condition).

As in the laboratory cohort we first aimed to assess molecular individuality and the reliability of the obtained data. Dimensionality reduction of the saliva samples obtained in the control condition revealed a clear separation between the first and second half of the competition season (Fig. 5a). This pattern indicates systematic shifts in the molecular saliva profiles over the course of a season, which may be caused by external influences throughout the year, or by an accumulation of load over the course of the season^28^.Despite the long sampling period of 16 months and the associated external variability, individual fingerprints remained recognizable in both the first and the second half of the season (Fig. 5a).

**Fig. 5.**
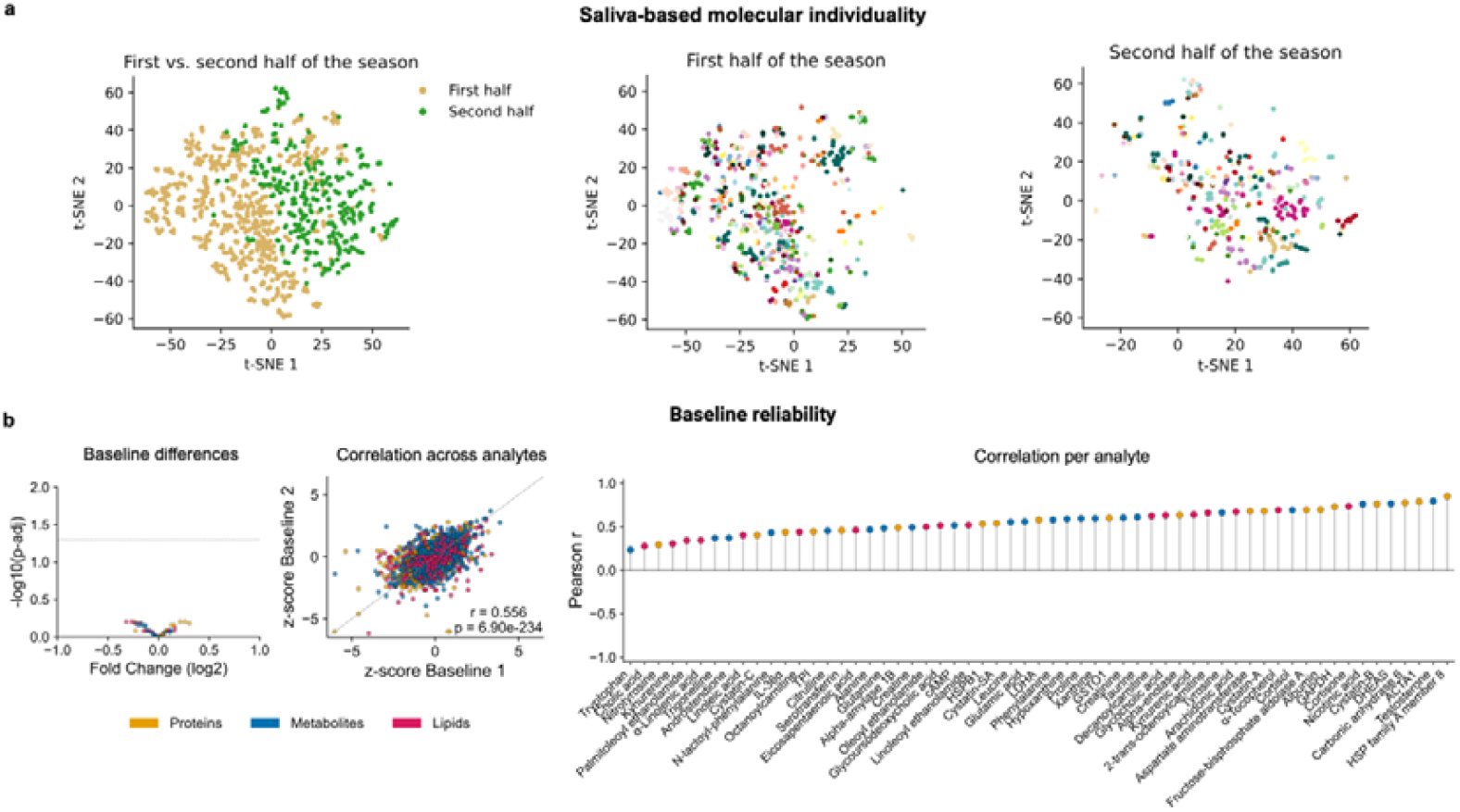
Real-world sampling confirms molecular individuality and high reliability of targeted saliva multi-omics. **a**, t-distributed stochastic neighbour embeddings (t-SNE) demonstrate that targeted saliva multi-omics can capture seasonal trends and molecular individuality. Samples are coloured by participant ID. N = 49 with 1011 samples and 59 analytes per sample. **b**, Baseline reliability was assessed by drawing the two samples in closest temporal proximity per participant ID. Two-sided, Benjamini-Hochberg-corrected, paired Student’s t-tests (left), overall correlation analyses (middle), and per-analyte correlations (right) were calculated, respectively. N = 49 with 98 samples and 59 analytes per sample.

To evaluate the reliability of the targeted multi-omics approach under real-world conditions, we examined the consistency of analyte measurements across two days in close temporal proximity. No analyte showed significant differences between the two days. Global correlation across all analytes was moderate (r = 0.56; p = 6.90 × 10^-234^), indicating considerable analytic stability despite increased biological and contextual variability. Furthermore, per-analyte correlations were moderate to high across the entire panel, underscoring the robustness of the measurements in a longitudinal real-world setting (Fig. 5c).

### The laboratory-based classifier framework is highly reproducible in a real-world setting

To assess whether the prediction model developed under controlled laboratory conditions can be transferred to real-world settings, we applied the same framework to longitudinal saliva samples. For model development, we created a balanced dataset (892 samples) to ensure an even distribution of exercise (match) and control samples per player. This dataset was used to train, test and fit the final model, while additional samples from the same players were set aside as held-out data for unseen internal validation (759 samples). Although conceptually comparable to the laboratory 24-hour prediction, sampling times in the real-world setting varied between 12 and 21 hours after exercise, resembling real-world challenges that introduce additional biological and contextual variance.

In the real-world setting, the most powerful model combination used LASSO-based feature pre-selection followed by the XGBoost classification, yielding a predictive ROC-AUC value of 0.84 and Bernoulli likelihood of 0.65 (Fig. 6a). In contrast to the laboratory setting, where only three features remained predictive 24 hours after exercise, the real-world model retained 54 of the 59 supplied features. This likely reflects the greater variability in sampling times, as different analytes become informative at different time intervals after exercise/control, as well as differences in exercise load between the standardised laboratory CPET and the more heterogeneous real-world football match setting. Nevertheless, predictive performance remained robust despite the non-standardised sampling window, indicating that indeed, our laboratory-based prediction framework can be transferred to real-world settings.

**Fig. 6.**
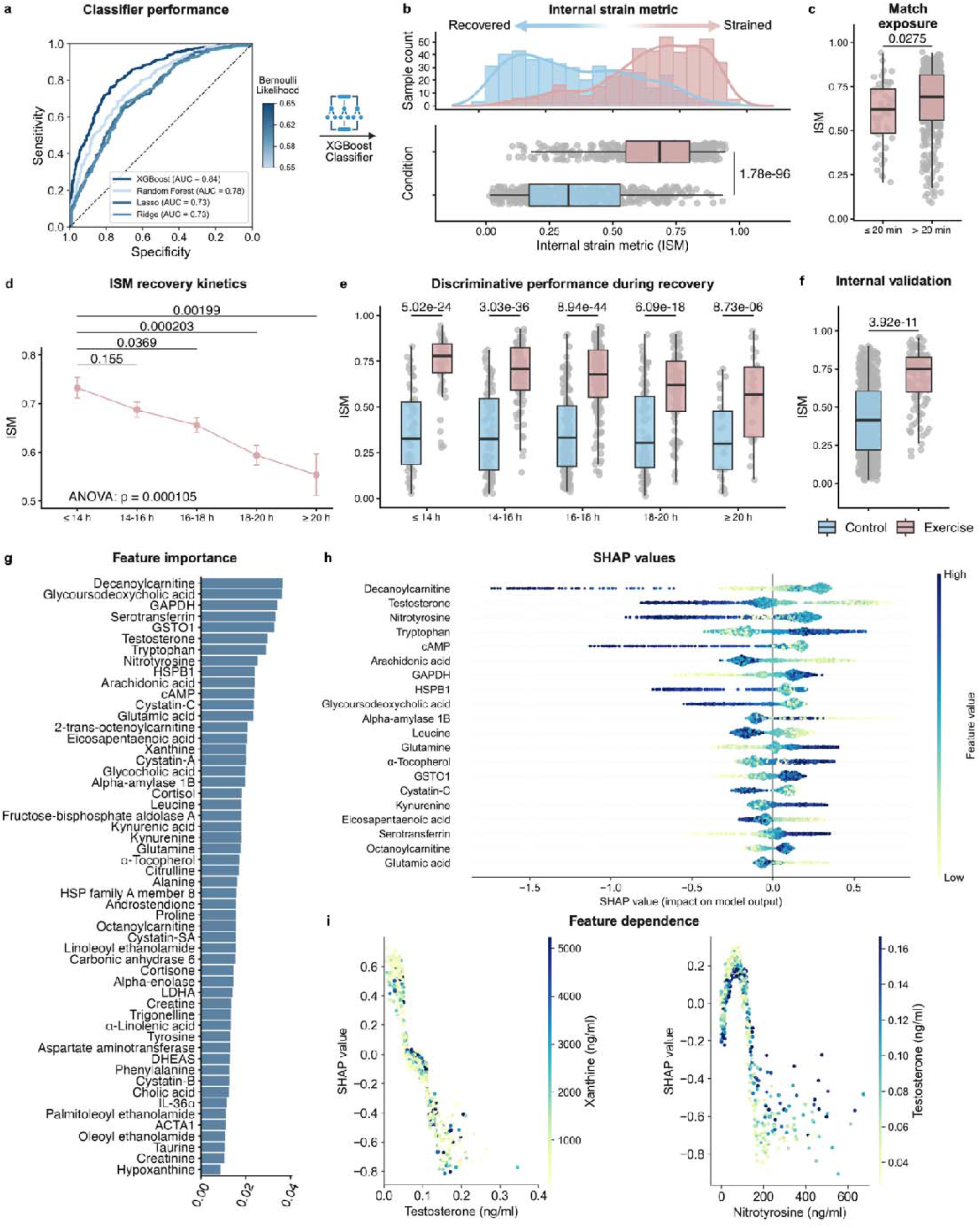
Real-world targeted saliva multi-omics captures physical exercise and recovery. **a**, Comparison of the predictive receiver operating characteristic area under the curve (ROC-AUC) across four models for classification of exercise vs. control the day after exercise. Predictive ROC-AUC curves are coloured by Bernoulli likelihood. **b**, An internal strain metric (ISM) that discriminates between exercise and control was derived from the prediction probabilities of the best-performing classifier (XGBoost). **c**, Comparison of the ISM between players that played ≤ 20 min versus > 20 min. **d**, The ISM decreases with progressing recovery time. Data depicted as mean ± SEM. **e**, The ISM discriminates between exercise and control across different recovery timeframes. Exercise and control samples were matched by player ID. **f**, Internal validation of the ISM on unseen data. **g**, Gain-based feature importance of XGBoost. **h**, SHapley Additive exPlanations (SHAP) values from XGBoost, showing the 20 most important features. **i**, SHAP dependence plots for Testosterone and Nitrotyrosine with features demonstrating highest interaction, respectively. The dots are color-coded according to the most influential interacting feature. All ISM values were compared using mixed ANOVAs implemented as a linear mixed-effects models. For post-hoc analysis of ISM values Holm-corrected pairwise comparisons between recovery timeframes and conditions were performed. Boxplots represent the 50^th^ (median), 25^th^, and 75^th^ percentiles (bounds of the box) with whiskers extending to 1.5 × the interquartile range. N = 49 with 892 samples and 59 analytes.

### Establishing the internal strain metric – a comprehensive measure of internal load

Being able to discriminate between exercise and control in a real-world setting, we next targeted our efforts at creating a practically relevant tool that delivers this discriminatory power to day-to-day decision-making in elite sports. In this context, a major unmet need for athlete monitoring is a comprehensive measurement of internal load, that reflects both exercise strain and the subsequent recovery process on a physiological level. To address this, we used the model probabilities predicted by the XGBoost classifier to define an internal strain metric (ISM), which reflects an individuals’ probability of having exercised on the day before. Conceptually, higher ISM values indicate incomplete recovery, and lower values indicate a return to the controlled (recovered) state. The ISM successfully discriminated between exercise and control samples (p = 1.78 × 10^-96^, Fig. 6b) and displayed higher values after short match exposure (≤ 20 min) compared to longer match exposure (> 20 min; p = 0.0275; Fig. 6c). This supports the assumption that the metric reflects the amount of match-related internal strain. To evaluate how the ISM changes during recovery from exercise, we divided the samples obtained on the day after exercise into 2-hour timeframe. Interestingly, the metric steadily decreased as recovery-time increased (p = 0.000105; Fig. 6d), suggesting that the ISM can additionally be harnessed to monitor recovery. Moreover, we compared the metric between exercise and control samples within each 2-hour timeframe, to determine whether the model retains discriminatory power across different sampling times. Across all time intervals, exercise samples consistently showed significantly higher ISM values than control samples (p = 0.0137; Fig. 6e), indicating that the metric reliably differentiates between exercise and recovery despite variable sampling times. This high discriminatory performance over an entire follow-up day creates strong applicability of our model for time-independent assessments of individual recovery trajectories.

To further evaluate robustness, we tested the final model on held-out samples collected at different time points during the season from the same individuals (Fig. 6f). The model achieved a predictive ROC-AUC of 0.82 and a Bernoulli likelihood of 0.6, demonstrating strong out-of-sample performance and good generalisability within the real-world setting.

Harnessing our interpretable machine learning approach, we next explored the underlying features of our ISM classifier. The XGBoost classifier, trained on all the data, revealed a broad distribution of gain-based importance across nearly all analytes (Fig. 6g). This pattern likely reflects continuous changes in the physiological state of the measured analytes over time after exercise, while circadian fluctuations - as previously detected in the laboratory control samples - further modulate analyte concentrations. Consequently, different features become informative at different sampling times, leading to a widely dispersed importance profile rather than a small group of dominant features.

SHAP analysis of the 20 most important features identified Decanoylcarnitine, Testosterone, and Nitrotyrosine as the top contributors (Fig. 6h). Similar to the laboratory setting, higher values of these analytes shifted predictions towards the control condition, whereas lower values increased the probability of being classified as exercise. Conversely, features such as Tryptophan, Glyceraldehyde 3-phosphate dehydrogenase (GAPDH), and Glutamine showed the opposite direction, with higher values indicating a higher probability of the exercise condition. Finally, dependency plots for Testosterone and Nitrotyrosine revealed clear non-linear relationships with the predicted probability, which likely contributes to the outperformance of the XGBoost classifier over other models (Fig. 6i).

Together, our real-world results show that a single-timepoint saliva sample can reliably discriminate between exercise and control conditions on the following day. With the probabilistic ISM introduced here, we suggest a comprehensive measure of internal load that depicts exercise strain and recovery and is based on non-invasive saliva sampling, which is easy to implement in different applied settings.

### DISCUSSION

Here, we demonstrated that high-frequency targeted saliva multi-omics is a reliable, non-invasive approach for capturing physical exercise and recovery across controlled laboratory and real-world settings. Our findings highlight saliva as a powerful, non-invasive biofluid for repeated sampling, enabling dense longitudinal profiling without the logistical and physiological challenges associated with blood-based assessments.

The identified molecular fingerprints are consistent with previous observations of molecular individuality in omics-based studies^8,25,29,30^. Through the precise and reproducible quantification of multiple analytes, our saliva-based targeted multi-omics analysis supports the establishment of individual reference ranges^23^ and facilitates integration with contextual information such as training load, GPS-data, or clinical metadata in real-world settings. This is crucial for interpreting dynamic physiological states, especially in long-term monitoring, where population-level thresholds often fail to capture meaningful changes within an individual.

Our dense 24-hour sampling revealed pronounced circadian modulation across proteins, metabolites, and lipids, providing the most extensive description of saliva-based circadian molecular dynamics to date and underscoring the importance of high temporal resolution when interpreting molecular measurements. Importantly, we observed circadian patterns for relevant exercise-responsive metabolites such as N-lactoyl-phenylalanine^24^ and Kynurenic acid^31^, that have been described primarily in blood and, when reported in saliva, have not previously been captured with such high temporal resolution. Of note, many biomarkers commonly used in opportunistic sampling strategies are strongly influenced by circadian rhythms^32,33^, making them difficult to interpret when considered separately. By resolving these temporal patterns, dense saliva profiling allows for a clearer distinction between circadian fluctuations and stress-related molecular changes, which is a requirement for accurate assessments of individual physiological states.

Building on this characterisation, machine learning-based modelling demonstrated that our targeted approach can be used to determine recent physical exercise based on individual time points, with predictive performance comparable to that reported for other biomarker-based monitoring studies^34,35^. Importantly, this analytical framework was translated to a longitudinal real-world cohort, where models retained robust discriminatory capacity despite increased biological and contextual variability. Together, these findings highlight the generalisability of saliva-based targeted multi-omics for monitoring dynamic physiological states beyond strictly controlled laboratory conditions.

Based on the results of the probabilistic model, we introduced the saliva-based ISM as a personalised measure of internal load. Conventional single biomarkers often reflect isolated biological processes and show limited robustness across individuals and training modalities, as exemplified by CK, which is commonly used to assess muscle damage^26^. Particularly in sports involving lower impact, lower eccentric loading, or little physical contact, such as cycling or swimming, the practical benefits of CK are limited. In contrast, the ISM integrates various biological signals, underscoring the suitability of our targeted multi-omics panel. This integrative design is consistent with approaches in health and immune monitoring^35–39^, and offers practical advantages for long-term decision-making, particularly in settings where frequent, non-invasive assessments are required.

Despite these promising results, our analysis is marked by several limitations that must be considered. Differences in cohort composition and contextual factors between laboratory and real-world settings limit direct comparability. Additionally comprehensive validation of our ISM against established strain parameters such as heart rate, energy expenditure or performance measures is urgently needed. Future studies involving broader populations and conditions, including different age groups, health statuses, and training loads are required to replicate and validate these findings. Moreover, extending high-frequency sampling beyond 24 hours (e.g. to 48 hours) would help to better capture longer-lasting recovery processes and improve our understanding of delayed molecular responses to physical exercise.

In conclusion, by combining a controlled trial with real-world applications, our framework – particularly the ISM – provides a blueprint for non-invasive, individualised physiological assessment across performance, health, and disease contexts. Overall, this establishes targeted saliva multi-omics as a robust and scalable approach for high-resolution monitoring of integrated physiological stress.

## Data availability

All data are available from the corresponding author on reasonable request.

## Ethics approval and informed consent

The studies were approved by the ethical committee at TU Dortmund University (approval number: GEKTUDO_2024-65 and GEKTUDO_2024-33) and were registered in the German Clinical Trials Register (registration numbers: DRKS00035227 and DRKS00034421). Informed consent was collected from all participants before data collection commenced. In the case of underage participants, parental consent was also obtained.

## METHODS

### Overview of cohorts

The data reported in this paper is derived from two cohorts, including a laboratory cohort of young female football players as well as a real-world cohort of elite male football players.

### Laboratory cohort

#### Study design

A randomised crossover study was conducted at TU Dortmund University after approval by the local ethics committee. Fourteen young female football players aged between 13 and 16 years were initially recruited, of whom two participants dropped out during the study due to acute illness, resulting in a final sample of 12 participants who were physically capable of cardiopulmonary exercise testing (CPET). After evaluation of exclusion criteria, written informed consent was obtained from all participants and their parents. Exclusion criteria comprised acute health complaints such as infections or injuries, chronic internal, orthopaedic, or psychological conditions that would contraindicate maximal CPET, short-term or long-term medication use except for the anti-baby pill, wearing a pacemaker, and suspected or confirmed pregnancy, which prevents bioimpedance analysis for body composition assessment. During the first visit, participant characteristics were collected, including age, use of hormonal contraception, pre-existing conditions, allergies, regular medication, alcohol and tobacco consumption. A physical examination and resting ECG recording were also conducted by a sports physician. Each participant then underwent laboratory testing in a sports medicine laboratory at TU Dortmund during an exercise and a control condition.

#### Study conditions

In the exercise condition CPET was performed on a cycle ergometer (ergoselect 5, ergoline GmbH) between 07:00 a.m. and 11:00 a.m. using breath-by-breath spirometry (Cortex Metalyzer 3B, CORTEX Biophysik GmbH). During the CPET, blood pressure and heart rate (Polar FS1C, Polar Electro) were recorded. The CPET protocol consisted of one-minute rest, a two-minute warm-up at 20 watts, a ramp protocol until volitional exhaustion, and a three-minute cool-down at 20 watts. Following the CPET, participants completed a 10-minute passive recovery period. A verification phase (VP) was then performed to confirm the attainment of maximal oxygen uptake, consisting of 1 minute at 60% of the previously attained maximal watts, followed by exercise at 110% of maximal watts performed to volitional exhaustion, and a subsequent 3-minute cool-down. In the control condition anthropometric data was collected, including height, weight, waist and hip circumference. Body composition was assessed using bioimpedance analysis (Seca mBCA 525, seca GmbH & Co. KG). Afterwards, participants were instructed to remain in a seated position for the same duration as in the exercise condition.

#### Randomization

The order of the study conditions was randomised by chronotype (exercise-control vs. control-exercise), and both conditions were separated by seven days to prevent potential carry-over effects. Participants were instructed to refrain from intensive physical activity, alcohol and caffeine consumption for 24 hours prior to each laboratory visit and arrive in an overnight-fasted state. At the end of each laboratory visit at TU Dortmund University, participants were provided with a standardised meal.

#### Measurement timepoints

Saliva samples were collected twice at baseline, immediately after the CPET, after the VP, and 15, 30, and 60 minutes after the VP. Following the samples under supervised conditions, participants consumed a standardised breakfast and self-collected saliva samples every two hours until 25 hours after the CPET. Additional samples were collected before going to bed and after waking up. Saliva samples were collected at identical timepoints in the control condition.

#### Real-world cohort

A real-world cohort of 50 elite male football players was monitored for a total of 16 months during their common training and match regimes. Saliva samples were drawn at the training facility multiple times per week and stored at 4°C until further analysis.

### Saliva sampling

#### Sample collection

Salivette® Cortisol (Sarstedt) tubes were used for saliva sampling according to the manufacturer’s instructions. The participants were instructed to rinse their mouths with water 10 minutes prior to sample collection with no further food or liquid intake permitted until sample collection. After sampling, water intake was allowed ad libitum. In brief, each participant placed a swab from a Salivette® tube in their mouth for one minute. The swab was then returned directly to the Salivette® tube without contact to any external surfaces. After sample collection all samples were frozen as quick as possible. During 24-hour sampling participants stored the samples at 4°C until the next day.

#### Liquid chromatography-tandem mass spectrometry

(LC-MS/MS) LC–MS/MS measurements were carried out in-house at the Biolyz Biomarker Platform. Depending on the method used, small molecules were extracted via protein precipitation, dilute-and-shoot or solid-phase extraction. The molecules were then separated based on exercise condition or reversed-phase C18 columns, operated under analytical or micro-flow conditions. Detection was performed by different triple quadrupole instruments in scheduled multiple reaction monitoring mode. For targeted proteomics, proteins were digested with trypsin using a bottom-up approach. The resulting surrogate peptides were resolved on micro-flow C18 LC and quantified by MRM with isotopically labelled standards. The platform delivered a linear dynamic range spanning four orders of magnitude, with limits of detection of < 1 pg/ml. System suitability was confirmed daily by using calibrants and pooled-matrix controls, resulting in average intra-run and inter-day coefficients of variation of ≤ 5% and ≤ 9%, respectively.

### Data processing

Before formal analysis of the obtained data, we implemented several processing steps aimed at ensuring analytic quality of the obtained data.

#### Laboratory cohort

Data obtained from the laboratory cohort was processed separately for assessment of baseline reliability, acute exercise effects, and 24-hour kinetics. The baseline dataset consisted of the samples obtained from the two baseline measurements of both intervention days. The acute exercise dataset consisted of the first baseline sample and the samples obtained immediately after the CPET, after the VP, and 15, 30, and 60 minutes after the VP. The 24-hour dataset consisted of all samples obtained by the participants without supervision. For the baseline dataset, we subdivided the data into a dataset for baseline reliability on day one, a dataset for baseline reliability on day two, and a dataset for inter-day reliability of baseline one. In each of these datasets we only retained analytes that were quantified above the lower limit of detection (LLOD) in ≥ 10 participants, resulting in 49 analytes for reliability assessment on day one (N = 11 due to one missing sample), 55 analytes for reliability assessment on day two (N = 12), and 54 analytes for inter-day reliability assessment (N = 12). For the acute exercise dataset and the 24-hour dataset we only retained analytes that were quantified above LLOD in ≥ 70 % of the samples and ≥ 10 participants in both conditions (exercise and control). This resulted in 54 analytes in the acute exercise dataset (N = 12) and 53 analytes in the 24-hour dataset (N = 12). The 24-hour dataset was manually supplemented with melatonin, which did not meet the predefined quality criteria but was nonetheless included to demonstrate physiological relevance, given its increasing concentrations during the evening hours. This resulted in a total of 53 analytes in the 24-hour dataset. Missing values and values quantified as LLOD were subsequently interpolated linearly per condition (i.e., exercise, control) and participant ID in the acute exercise dataset and the 24-hour dataset to retain individual analyte kinetics. In case of missing values or LLODs at the margins of the implemented measurement timepoints constant extrapolation was applied.

#### Real-world cohort

Data obtained from the real-world cohort was processed by only retaining samples of players that played in a match the day prior (= exercise) or that did not play in a match or train for ≥ 3 days (= control). The goalkeeper was excluded from the dataset due to different physical demands during a match, which resulted in a dataset of 49 players. Next, we only retained analytes that were quantified in ≥ 70 % of the samples and imputed missing values and values quantified as LLOD separately for exercise and control using the `missForest` package. This resulted in a dataset containing 1,651 samples and 59 analytes per sample. We then created a separate dataset containing in-season control samples for evaluation of baseline reliability, which comprised 1,011 samples. Additionally, we created a balanced dataset containing equal numbers of exercise and control samples per player ID (892 samples). This procedure ensured equal class representation within individuals while allowing between-participant differences in the total number of observations. Additional samples from the same participants, not included in the balanced dataset, were retained for unseen internal validation (759 held-out samples).

### Statistical analysis

All statistical analyses were conducted in Python^40^ (version 3.13.9) or R^41^ (version 4.5.1.) at a significance level of 5⍰%. In R the `dplyr` and `tidyr` package were used for data wrangling and the `ggplot2` and `ggpubr` package were used for visualization. In Python `Pandas` and `NumPy` were used for data wrangling and `Matplotlib` was used for visualisation.

#### Saliva-based molecular individuality

T-distributed stochastic neighbour embeddings (t-SNE) were applied to visualise molecular individuality of saliva-based multi-omics data using z-transformed and log_2_-transformed concentrations. Analyses were performed in Python using `sklearn.manifold.TSNE` and visualized with `Matplotlib`. For the samples obtained from the laboratory cohort we employed a two-dimensional reduction (n_components = 2) with a fixed random state of 50 and a maximum of 9,000 iterations. Perplexity was set to 13 for t-SNE of control samples and 18 for t-SNE of baseline samples. For the samples obtained in the real-world cohort, we employed a two-dimensional reduction with a fixed random state of 50 and a perplexity of 10.

#### Baseline reliability

Baseline reliability was assessed via three different approaches using log_2_-transformed concentrations. First, baselines concentrations were compared using two-sided, Benjamini-Hochberg-corrected, paired Student’s t-tests. Second, Pearson’s correlation coefficients were calculated across all analytes and samples. Third, Pearson’s correlation coefficients were calculated for each analyte. For correlation analyses z-transformed values were used. To obtain baseline samples from the real-world cohort we selected two consecutive sampling days per played ID based on highest temporal proximity.

#### Analysis of 24-hour kinetics

24-hour kinetics were analysed using all control samples from the 24-hour dataset. Bayesian Additive Models (BAMs) were calculated in R based on interpolated concentrations. Each analyte was modelled independently over time with k = 10 using the `mgcv` package. Melatonin was modelled independently using raw concentrations with k = 5, due to lower data density, as it was predominantly quantified during evening hours in accordance with physiological fluctuations.

#### Inter-individual variability of analytes

Inter-individual variability was evaluated by calculating coefficients of variation, which are defined as the ratio of the standard deviation to the mean. Coefficients of variation were calculated separately for each analyte, condition, and measurement time point and then summarized for each omics layer.

#### Analysis of exercise effects

Linear mixed models (LMM) were employed using log_2_- and z-transformed concentrations to examine changes in analytes over time and time × condition interaction effects in the acute exercise dataset and in the 24-hour dataset. The LLM included fixed effects for time point, condition, and the interaction between both as well as a random intercept for participant ID. Models were implemented using the `lme4` and `lmerTest` package in R. Based on the LMM results, analysis of variance was performed. In case of significant time × condition interaction effects, we implemented post-hoc analyses using the `emmeans` package to identify time effects within each condition and condition effects within each time point. For post-hoc analysis two-sided, Bonferroni-corrected, paired Student’s t-tests were performed. A heatmap was created to visualize acute analyte changes over time separated by condition. For each analyte, condition, and timepoint, log_2_ fold changes from baseline were calculated. To identify similar kinetics across analytes within each omics layer we applied hierarchical clustering. Euclidean distance and dendrograms were calculated in Python with `scipy.cluster.hierarchy` and `scipy.spatial.distance.`

### Prediction analyses

#### Overview of prediction settings

A total of three prediction settings were considered using saliva-based analytes. The first setting included saliva samples from immediately after exercise in the laboratory cohort. The second setting included saliva samples from 23 to 25 hours after exercise in the laboratory cohort. Since data from one participant was unavailable at this time point, only eleven participants were included in this model. The third setting included saliva samples from the balanced dataset obtained in the real-world cohort (see above for data processing and dataset curation). Across all settings, the regarded outcome variable was binary (exercise vs. control). Saliva analytes served as input features for the prediction models, with their number varying slightly across settings. Specifically, 52 analytes were included in the first setting, 54 in the second setting, and 59 in the third (real-world) setting. All features were log_2_-transformed and subsequently z-score standardised prior to model fitting.

Given the repeated-measures design, different mixed-effects models with participant-specific random intercepts were explored in preliminary analyses. However, across all settings, random-effects variances were small to moderate relative to the fixed effects, indicating that the features explained most of the outcome variability. Therefore, all subsequent analyses did not explicitly account for the repeated-measures design.

#### Model development and comparison

Model comparison was performed using a nested cross-validation (CV) framework that explicitly separated feature selection, hyperparameter tuning, and model evaluation. An outer leave-one-subject-out cross-validation (LOSO-CV) to assess the predictive performance of different statistical and machine learning approaches was applied, such that in each outer iteration all samples from one participant were excluded from model fitting and used solely for evaluation, while the remaining participants constituted the training set. Within each outer training set, all feature selection and hyperparameter optimisation steps were conducted using five-fold CV. This inner CV scheme was identical across all models and settings.

Two alternative feature selection strategies were evaluated and contrasted within this framework. In the first strategy, feature selection was performed using LASSO-penalised logistic regression. The penalty parameter was optimised via five-fold CV using receiver operating characteristic area under the curve (ROC-AUC) as the optimisation criterion. Based on the optimised LASSO model, all features with non-zero coefficients were selected and retained for subsequent model training. In the second strategy, feature selection was performed using random forest models, again optimised via five-fold CV and using ROC-AUC as the optimisation criterion. For each outer fold, the top 20 features ranked by feature importance were selected. Following the feature selection step, four classification models were compared (solely based on the selected features): LASSO-penalised logistic regression^42,43^, ridge-penalised logistic regression^44^, random forest^45^, and extreme gradient-boosted decision trees (XGBoost)^46^. For each classifier, model-specific hyperparameters were optimised, again using five-fold CV within the outer training set. The resulting optimised models were then evaluated on the data excluded in the corresponding outer LOSO iteration.

Predicted probabilities from all outer folds were pooled and averaged to compute overall performance metrics. Model performance was assessed using the predictive ROC-AUC. In addition, the predictive Bernoulli likelihood was computed based on the predicted probabilities as a complementary measure of probabilistic calibration, defined as the average probability assigned to the true class label across all samples. The combination of feature-selection strategy and classifier yielding the highest ROC-AUC and Bernoulli likelihood was selected as the winning model. Finally, for completeness, we also regarded the Brier score and classification rate.

### Definition of the saliva-based internal strain metric

From the prediction analysis of saliva samples obtained in the real-world cohort, we derived a quantitative measure of internal load, which we termed “Internal Strain Metric (ISM)”, defined as the probability of having exercised on the previous day based on the best-performing classifier (XGBoost). ISM values were derived exclusively from LOSO-CV predictions to avoid optimistic bias. For comparisons between exercise and control and between different match durations mixed ANOVAs were implemented as linear mixed-effects models with condition as a fixed effect and player ID as a random intercept. For analysis of ISM values across different recovery timeframes a linear mixed effects model was fitted and Holm-corrected pairwise comparisons between recovery timeframes were performed. Additionally, a separate model was fitted with recovery timeframe, condition, and their interaction as fixed effects and player ID as random intercept. Holm-corrected post-hoc comparisons of conditions within each recovery window were performed.

### Final model training and evaluation

For each setting, the best-performing combination of feature selection method and classifier was retrained on the full dataset using five-fold CV for hyperparameter optimization. Coefficient estimates (corresponding to standardised features) for the regression approaches and gain-based feature importance measures were extracted from the final models to support biological interpretation. The optimal model configurations differed between settings: random forest feature selection combined with ridge-penalised logistic regression in the first setting, and LASSO-penalised logistic regression feature selection combined with XGBoost in second and third settings. In the third setting only, the final model was additionally evaluated on the held-out samples that were not included in model development. Although originating from the same participants, these samples had not been used during training or cross-validation. Model performance on the held-out dataset was evaluated using ROC-AUC and predictive Bernoulli likelihood, complemented by the Brier score and classification rate.

