## Supplementary Table 1 for "Targeted saliva multi-omics is a reliable, non-invasive method to capture physiological stress and recovery"

**Table 1** Participant characteristics (laboratory setting)

|  | | **Mean ± standard deviation [min; max]** |
| --- | --- | --- |
| **Demographic and anthropometric characteristics** | | |
|  | Sex, f | 12 |
|  | Age, yr | 15.17 ± 1.03 [13; 16] |
|  | Height, cm | 166.58± 5.18 [173.0; 158.0] |
|  | Weight, kg | 56.95± 6.83 [46.3; 64.3] |
|  | BMI, kg/m^2^ | 20.20 ± 2.1 [17.21; 23.62] |
|  | Relative fat mass, % | 23.05± 5.53 [16.46; 31.8] |
|  | Fat-free mass, % | 43.27 ± 3.22 [36.57; 48.34] |
|  | Skeletal muscle mass, % | 20.17 ± 2.12 [15.99; 22.6] |
| **Cardiopulmonary exercise test** | | |
|  | V̇O_2peak,_ mL/min/kg | 44.17 ± 5.39 [35.0; 52.0] |
|  | Maximal heart rate, bpm | 185.5± 21.63 [122; 208] |
|  | Absolute PPO, W | 199.33± 32.26 [149; 252] |
|  | Relative PPO, W/kg | 3.54 ± 0.38 [2.9; 4.1] |
| **Cardiopulmonary verification phase** | |  |
| V̇O_2peak,_ mL/min/kg | | 43.83 ± 5.67 [33.55; 52.2] |
| Maximal heart rate, bpm | | 187 ± 12.41 [163; 202] |

BMI, body mass index; V̇O_2peak_, peak oxygen uptake, PPO, peak power output. Due to technical reasons heart rate data was only recorded from 10 participants in the verification phase.
